# Multi-omics analysis reveals aspirin is associated with reduced risk of Alzheimer’s disease

**DOI:** 10.1101/2025.04.07.25325038

**Authors:** Monika E. Grabowska, Avi U. Vaidya, Xue Zhong, Chris Guardo, Alyson L. Dickson, Mojgan Babanejad, Chao Yan, Yi Xin, Sergio Mundo, Josh F. Peterson, QiPing Feng, James Eaton, Zhexing Wen, Bingshan Li, Wei-Qi Wei

**Author notes:** Corresponding author: Wei-Qi Wei, MD, PhD Associate Professor, Department of Biomedical Informatics Vanderbilt University Medical Center, Address: Suite 1500, 2525 West End Ave, Nashville, TN, USA, 37203.

## Abstract

The urgent need for safe and effective therapies for Alzheimer’s disease (AD) has spurred a growing interest in repurposing existing drugs to treat or prevent AD. In this study, we combined multi-omics and clinical data to investigate possible repurposing opportunities for AD. We performed transcriptome-wide association studies (TWAS) to construct gene expression signatures of AD from publicly available GWAS summary statistics, using both transcriptome prediction models for 49 tissues from the Genotype-Tissue Expression (GTEx) project and microglia-specific models trained on eQTL data from the Microglia Genomic Atlas (MiGA). We then identified compounds capable of reversing the AD-associated changes in gene expression observed in these signatures by querying the Connectivity Map (CMap) drug perturbation database. Out of >2,000 small-molecule compounds in CMap, aspirin emerged as the most promising AD repurposing candidate. To investigate the longitudinal effects of aspirin use on AD, we collected drug exposure and AD coded diagnoses from three independent sources of real-world data: electronic health records (EHRs) from Vanderbilt University Medical Center (VUMC) and the National Institutes of Health *All of Us* Research Program, along with national healthcare claims from the MarketScan Research Databases. In meta-analysis of EHR data from VUMC and *All of Us*, we found that aspirin use before age 65 was associated with decreased risk of incident AD (hazard ratio=0.76, 95% confidence interval [CI]: 0.64-0.89, *P*=0.001). Consistent with the findings utilizing EHR data, analysis of claims data from MarketScan revealed significantly lower odds of aspirin exposure among AD cases compared to matched controls (odds ratio=0.32, 95% CI: 0.28-0.38, *P*<0.001). Our results demonstrate the value of integrating genetic and clinical data for drug repurposing studies and highlight aspirin as a promising repurposing candidate for AD, warranting further investigation in clinical trials.

---

Alzheimer’s disease (AD) remains a major public health concern, with current treatment options offering only modest benefits. Although over 32 million people are diagnosed with AD worldwide, estimates suggest that an additional 384 million are affected by early-stage (preclinical and prodromal) AD^1^, indicating that the true burden of AD is likely underestimated and highlighting the critical need for preventive strategies that can address the disease before symptoms emerge. Drug development for AD has been expensive, time-consuming, and prone to failure based on lack of efficacy or serious adverse effects^2^. As of 2025, only eight drugs have been approved for the treatment of AD in the United States: three amyloid-targeting disease-modifying therapies for mild AD (one of which has been discontinued) and five drugs for alleviating cognitive symptoms^3–5^. In this context, drug repurposing has emerged as a cost-effective supplemental strategy to evaluate whether any existing drugs may effectively treat or prevent AD. Beyond its potential to accelerate drug development timelines and reduce costs, drug repurposing allows for the exploration of novel and diverse disease mechanisms through the actions of well-established drugs with known safety profiles, making it an attractive strategy for expanding the therapeutic options available for AD.

One popular computational drug repurposing approach involves the identification of compounds whose transcriptomic profiles negatively correlate with a gene expression signature for a disease of interest. The underlying hypothesis is that such drugs could mitigate or reverse disease-associated changes in gene expression, thereby preventing the disease or limiting its progression^6–10^. While early studies primarily used differential gene expression analysis to define disease signatures, transcriptome-wide association study (TWAS) methodologies have emerged as a promising alternative. TWAS integrates expression quantitative trait loci (eQTL) and genome-wide association study (GWAS) data to identify genes whose expression levels are significantly associated with disease risk^11^, offering unique insight into candidate causal genes and potential therapeutic targets. Additionally, TWAS enables mapping of disease-associated gene expression patterns across multiple tissues, increasing the biological relevance and applicability of identified targets. Given evidence that drugs targeting genetically supported genes are approximately 2.6 times more likely to succeed in clinical development^12^, TWAS-based strategies hold promise for uncovering effective drug repurposing candidates, and have already been applied to conditions such as endometrial cancer, hypertension, hyperlipidemia, and nicotine addiction^13–15^.

The growing availability of large-scale public databases has further increased the scope and efficiency of computational drug repurposing via gene expression signature reversal, facilitating the development of repurposing pipelines integrating diverse datasets to systematically identify potential therapeutic candidates^14,16–18^. For TWAS-based approaches, eQTL datasets have provided a critical foundation for understanding gene expression regulation across diverse tissues and cell types, helping to elucidate the molecular mechanisms underlying complex disease. Landmark projects like the Genotype-Tissue Expression (GTEx) project^19^, which includes eQTL data for 49 different human tissues, have been instrumental in revealing tissue-specific patterns of gene expression. Drug perturbation databases, such as the Integrative Library of Integrated Network-based Cellular Signatures (iLINCS)^20^ and the Connectivity Map (CMap)^21^, have further advanced repurposing efforts by enabling direct comparison of drug-induced transcriptional profiles with disease signatures. In recent years, the incorporation of large clinical datasets has added another layer of validation to drug repurposing workflows^22,23^. Longitudinal analyses leveraging electronic health records (EHRs) and healthcare claims have sought to substantiate the efficacy and safety of proposed repurposing candidates using real-world patient data.

In this study, we integrate genomic, transcriptomic, and clinical data to generate and validate drug repurposing hypotheses for AD. We first identified AD risk genes using TWAS, integrating publicly available GWAS summary statistics and eQTL data from both bulk tissues and primary microglia. We then used the identified risk genes to construct cross-tissue and microglia-specific transcriptomic signatures of AD. Next, we screened the CMap drug perturbation database for compounds inducing opposing changes in gene expression, which suggested aspirin as a potential repurposing candidate for AD prevention. We then conducted clinical validation studies using longitudinal EHR data from Vanderbilt University Medical Center and the National Institutes of Health *All of Us* Research Program and health insurance claims data from the MarketScan Research Databases to investigate the association between aspirin and AD.

## Results

### Cross-tissue and microglial transcriptional signatures of AD

We applied a multi-omics approach to generate drug repurposing hypotheses for AD (**Fig. 1**). We performed transcriptome-wide association study (TWAS) using S-PrediXcan^24^ and S-MultiXcan^25^ to identify candidate risk genes for AD. Using pre-trained transcriptome prediction models for 49 tissues from the Genotype-Tissue Expression (GTEx) project and publicly available AD GWAS summary statistics^26^, we conducted three S-MultiXcan analyses balancing tissue specificity and statistical power: (1) a brain-specific analysis combining gene expression predictions for the 13 GTEx brain tissues, (2) an AD-relevant tissue analysis combining predictions for the 13 GTEx brain tissues along with four peripheral tissues previously related to AD (whole blood, spleen, and sun-exposed and unexposed skin tissues)^27,28^, and (3) a non-specific analysis combining predictions for all 49 GTEx tissues to maximize statistical power. In each S-MultiXcan analysis, AD risk genes were identified using a Bonferroni-corrected significance threshold (*P*<0.05/number of tested gene associations). We then developed three distinct AD gene expression signatures—for brain tissue, AD-relevant tissue, and all tissues combined—by selecting the risk genes with concordant effect directions (based on the sign of S-PrediXcan *Z* scores) across at least two-thirds of the tissues with available results analyzed in each respective S-MultiXcan analysis (13 brain tissues, 17 AD-relevant tissues, and all 49 GTEx tissues). We required this tissue-wide consensus because several genes with statistically significant S-MultiXcan results exhibited opposing effects across different tissues, producing near-zero effect estimates when averaged in the S-MultiXcan analysis (e.g., *APOE* in the all-tissue analysis, as illustrated in **Supplementary Fig. 1**). The AD risk genes identified in each S-MultiXcan analysis are provided in **Supplementary Tables 1**-**3**, along with the tissue-specific S-PrediXcan results for these genes, the consensus direction of effect, and the mean *Z* score computed across the tissues with the consensus effect direction.

**Fig. 1:**
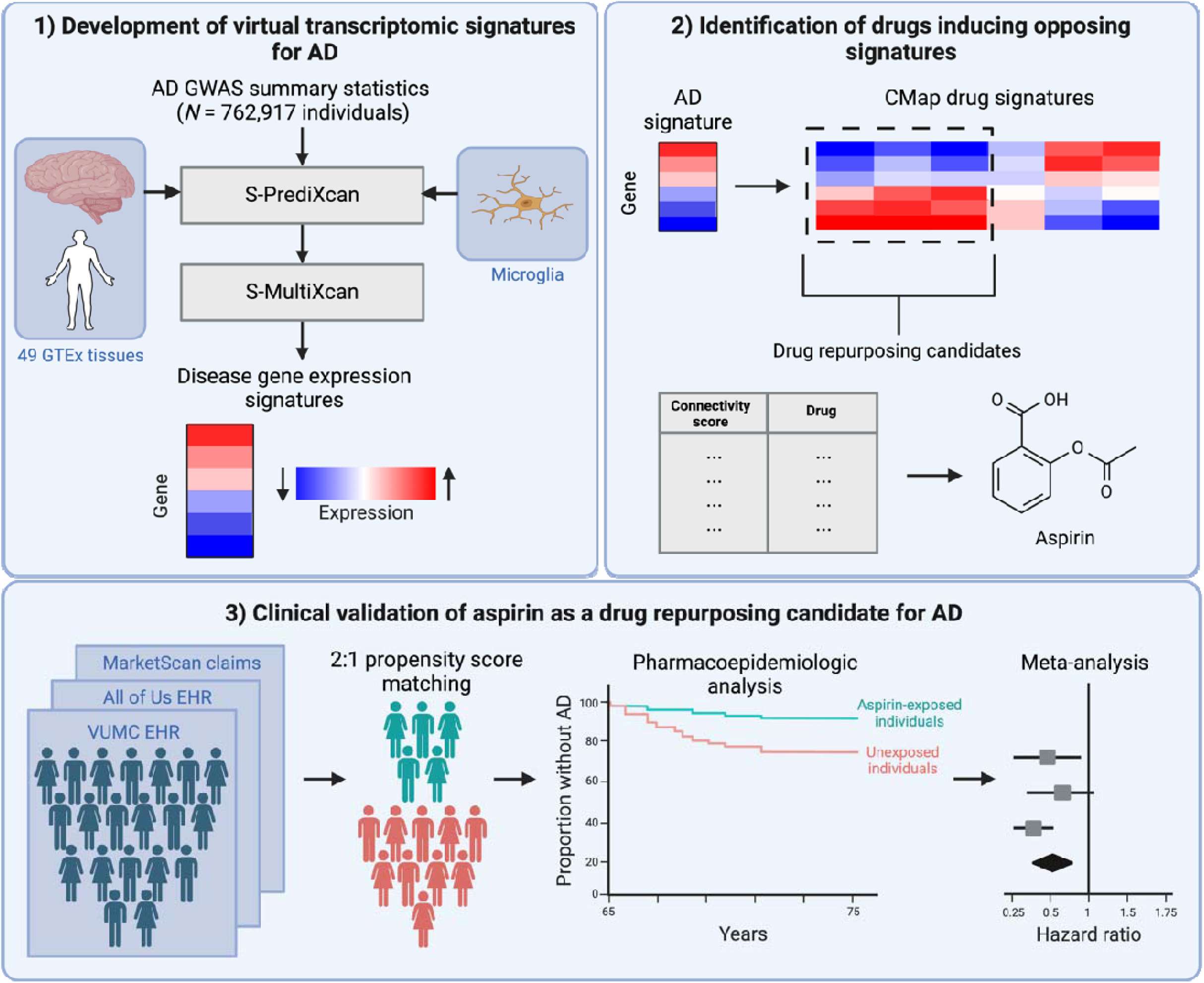
Study design and workflow. (1) Cross-tissue and microglia-specific gene expression changes associated with AD were imputed from AD GWAS summary statistics using S-PrediXcan and S-MultiXcan. The identified AD risk genes were used to construct disease gene expression signatures for AD. (2) The AD gene expression signatures were used to query the CMap drug perturbation database to identify drugs inducing opposing changes in gene expression. CMap queries identified aspirin as a potential drug repurposing candidate for AD. (3) Clinical validation studies investigating the effect of aspirin use on AD in humans were performed using longitudinal EHR data from VUMC, with independent replication using EHR data from the NIH *All of Us* Research Program database and healthcare claims data from the MarketScan Research Databases. AD Alzheimer’s disease, GWAS genome-wide association study, GTEx Genotype-Tissue Expression, CMap Connectivity Map, EHR electronic health record, VUMC Vanderbilt University Medical Center, NIH National Institutes of Health.

The final brain-tissue AD transcriptomic signature contained 72 genes (37 positively associated with AD risk, 35 inversely associated with AD risk), while the AD-relevant tissue signature contained 78 genes (41 positively, 37 inversely associated) and the all-tissue signature contained 79 genes (40 positively, 39 inversely associated) (**Fig. 2a**). Among the three signatures, 43 shared AD risk genes were identified (21 positively, 22 inversely associated).

**Fig. 2:**
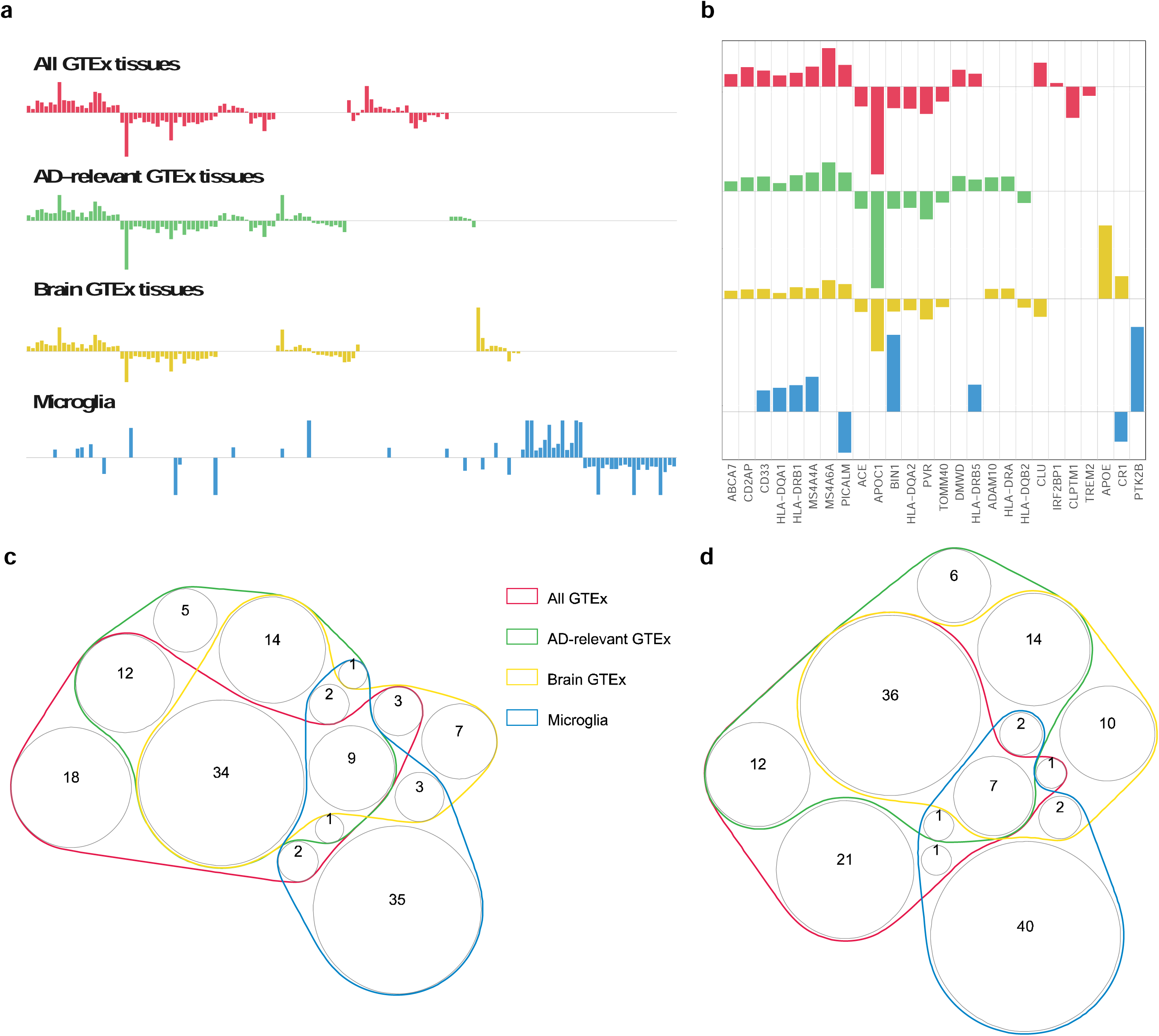
Transcriptomic signatures of AD. **a)** Bar plots comparing the four AD gene expression signatures used to query CMap. **b)** Bar plots highlighting predicted expression levels for genes with previously demonstrated relevance to AD. **c)** Venn diagram comparing the shared and unique AD risk genes in the GTEx and microglia transcriptomic signatures, irrespective of effect direction. **d)** Venn diagram comparing the shared and unique AD risk genes with concordant effect directionality in the GTEx and microglia transcriptomic signatures. AD Alzheimer’s disease, GTEx Genotype-Tissue Expression, CMap Connectivity Map.

To address the challenges associated with interpreting cell type-specific gene expression changes in heterogeneous bulk tissues in AD, we also trained custom transcriptome prediction models for four microglia-containing brain regions (medial frontal gyrus, superior temporal gyrus, subventricular zone, and thalamus) using eQTL summary statistics from the Microglia Genomic Atlas (MiGA)^29^. We then ran S-PrediXcan and S-MultiXcan analyses to identify AD risk-associated genes specific to microglia. However, S-MultiXcan produced valid association statistics for <20% (1,835/9,687) of the genes tested; thus, we applied the Generalized Berk-Jones (GBJ) test^30^ to combine S-PrediXcan predictions from the four brain regions. Because the GBJ test aggregates association statistics without indicating the direction of effect, we used the AD risk genes exhibiting consistent effect directions across all brain regions with available results to construct a single microglial AD gene expression signature. The GBJ results are provided in **Supplementary Table 4**.

Our microglial AD signature included 53 genes (25 positively associated with AD risk, 28 inversely associated with AD risk) (**Fig. 2a**). Across all four signatures, we identified several known AD risk-associated genes^27,31–59^, including *ABCA7*^31,32^, *BIN1*^40,47^, *CD33*^34–37^, *CLU*^41^, *CR1*^41,58^, *CD2AP*^33,34^, *MS4A6A*^41^, and *PICALM*^41,42^ (**Fig. 2b**). A comparison of the four AD transcriptomic signatures with all genes labeled is shown in **Supplementary Figure 2**. Only 18 of the genes included in the microglial AD signature overlapped with at least one of the three GTEx signatures (**Fig. 2c**), although four of these overlapping genes showed discordant associations with AD risk in the microglia and GTEx signatures (**Fig. 2d**).

### Aspirin reverses gene expression changes associated with AD

We then uploaded the risk genes comprising each AD gene expression signature into the CMap drug perturbation database^60^ to identify drugs with the potential to reverse disease-associated transcriptomic changes. CMap contains transcriptional profiles for over 8,000 small-molecule compounds and genetic modifications tested in multiple human cell lines, with the ability to rank perturbagen profiles based on similarity to a given disease signature through calculation of a connectivity score. Consistent with previous drug repurposing studies^14,17^, we defined potential repurposing candidates as compounds with negative computed connectivity (i.e., connectivity score<0) to our input AD signature. Out of 2,428 small-molecule compounds in CMap, 590 (24.3%) were negatively correlated with the GTEx brain-tissue AD signature, 709 (29.2%) were negatively correlated with the AD-relevant tissue disease signature, and 688 (28.3%) were negatively correlated with the all-tissue disease signature, representing possible AD drug repurposing candidates (**Supplementary Tables 5**-**7**). Of these, 218 drugs exhibited negative connectivity with all three GTEx AD signatures, including mycophenolate, fluticasone, sirolimus, sertraline, clozapine, and losartan, although the connectivity magnitude was subject to substantial variability (e.g., connectivity scores of -72.2, -7.6, and -0.6 for mycophenolate with the GTEx brain-tissue, AD-relevant tissue, and all-tissue signatures, respectively). Additionally, 794 (32.7%) small-molecule compounds were negatively correlated with the microglia-specific AD signature (**Supplementary Table 8**). The top ten AD repurposing candidates identified using each of the four AD gene expression signatures are shown in **Table 1**. Only two preclinical drugs were in all four lists of repurposing candidates: prostratin, an herbal protein kinase C activator, and BX-795, an IκB kinase (IKK) inhibitor.

**Table 1.** Top ten AD repurposing candidates identified in CMap queries using GTEx-derived and microglial disease signatures. Drugs approved by the United States Food and Drug Administration are marked with an asterisk and their clinical indications are provided in parentheses.

| GTEx brain tissues | AD-relevant GTEx tissues | All GTEx tissues | Microglia |
| --- | --- | --- | --- |
| mupirocin*<br>(impetigo and other uncomplicated bacterial skin infections) | CAY-10618 | indinavir*<br>(human immunodeficiency virus) | anisomycin |
| mycophenolic-acid*<br>(prophylaxis of organ rejection, autoimmune disease) | bufalin | isogedunin | emetine |
| BX-795 | vidarabine*<br>(herpes simplex keratitis) | deforolimus | homoharringtonine*<br>(chronic myeloid leukemia) |
| fluticasone*<br>(asthma, allergic rhinitis, and certain inflammatory skin conditions) | PP-30 | phorbol-12-myristate-13-acetate | narciclasine |
| prostratin | ivermectin*<br>(parasitic infections) | U-46619 | digitoxigenin |
| PD-123319 | rhodomyrtosin-b | BX-795 | troxipide |
| phenylbutyrate*<br>(urea cycle disorders) | pentylentetrazol | aspirin*<br>(pain, fever, primary and secondary cardiovascular disease prevention, rheumatoid arthritis) | pyrvinium-pamoate |
| desoxypeganine | ingenol*<br>(actinic keratosis) | pirinixic-acid | roscovitine |
| SA-63133 | prostratin | ON-01910 | cycloheximide |
| indinavir*<br>(human immunodeficiency virus) | salubrial | praziquantel*<br>(parasitic infections) | isoliquiritigenin |

Aspirin was identified as one of the top ten drugs with negative connectivity to the all-tissue GTEx AD signature (connectivity score = -69.96), a finding of particular interest since aspirin is a relatively well-tolerated drug taken by many patients, making it a favorable candidate for repurposing. This signal was replicated using the disease gene expression signature derived from the AD-relevant GTEx tissues (connectivity score = -17.89) and the microglial AD gene expression signature (connectivity score = -30.88), providing additional support for aspirin as a promising candidate, although we did not observe any signal for aspirin with the GTEx brain-tissue signature (connectivity score = 0). Given the large number of preclinical drugs with unknown side effects (e.g., prostratin and BX-795) and approved drugs with limited clinical use cases (e.g., indinavir, ivermectin, mycophenolate) returned among the CMap queries, we focused our subsequent analyses on evaluating aspirin’s potential as a drug repurposing candidate for AD.

### Aspirin exposure is associated with reduced risk of incident AD

We assessed our genetics-informed drug repurposing hypothesis by performing clinical validation studies investigating the relationship between aspirin use and AD diagnosis, leveraging real-world data from three large data sources: (1) VUMC’s de-identified EHR database, (2) the NIH *All of Us* Research Program database, and (3) national healthcare claims data from the MarketScan Research Databases. In the EHR validation studies, we used a retrospective cohort study design to compare AD incidence after age 65 between aspirin-exposed patients and a propensity score-matched cohort of unexposed patients. In contrast, due to the shorter observation periods within the claims data, which restricted our ability to observe AD diagnoses among individuals exposed to aspirin before age 65, we used a case-control study design to compare the odds of previous aspirin exposure in patients with AD versus propensity score-matched controls without AD. This approach allowed us to capture patients with both aspirin use and AD in the MarketScan data, which we were unable to do with the original retrospective cohort study design. Descriptive characteristics for the matched cohorts from the EHR-based validation studies are shown in **Table 2**. The characteristics of the matched MarketScan claims-based cohort are provided in **Supplementary Table 9.** Information on AD outcomes in all three datasets is provided separately in **Supplementary Table 10**.

**Table 2.** Description of matched patient cohorts used in EHR validation studies.

| Clinical characteristics | VUMC<br>(N = 19,413) |  | All of Us<br>(N = 1,995) |  |
| --- | --- | --- | --- | --- |
|  | Exposed | Unexposed | Exposed | Unexposed |
| **Aspirin** |  |  |  |  |
| N | 6,656 | 12,757 | 666 | 1,329 |
| Mean age at last follow-up (s.d.) | 77.8<br>(2.8) | 77.9<br>(2.8) | 77.3<br>(2.4) | 77.3<br>(2.4) |
| **Sex (%)** |  |  |  |  |
| Female | 49.8 | 51.8 | 45.6 | 45.7 |
| Male | 50.2 | 48.2 | 52.4 | 52.5 |
| **Race (%)** |  |  |  |  |
| White | 92.3 | 92.2 | 77.3 | 77.4 |
| Black | 6.8 | 6.8 | 7.1 | 7.3 |
| Other | 0.9 | 1.0 | 15.6 | 15.3 |
| **Baseline comorbidities (%)** |  |  |  |  |
| Cardiovascular disease | 25.1 | 21.2 | 28.5 | 28.4 |
| Cerebrovascular disease | 6.0 | 5.6 | 8.1 | 7.5 |
| Rheumatoid arthritis | 3.6 | 4.1 | 4.1 | 4.1 |

Using EHR data from VUMC, we observed that aspirin use before age 65 was associated with a significantly reduced risk of incident AD after age 65 (hazard ratio [HR]=0.77, 95% confidence interval [CI]: 0.65-0.91, *P*=0.003). We also investigated whether the protective effect of aspirin differed by dose, with high-dose use defined as aspirin doses ≥325mg/day and low-dose use defined as doses ≤81mg/day^61^. We observed a protective effect for individuals exposed to both high-dose aspirin (HR=0.63, 95% CI: 0.45-0.90, *P*=0.01) and low-dose aspirin (HR=0.82, 95% CI: 0.68-0.99, *P*=0.04) relative to no use, although the difference between high- and low-dose aspirin was not statistically significant (HR=0.77, *P*=0.19).

As patients prescribed high-dose aspirin often have more severe cardiovascular and cerebrovascular conditions compared to those on low-dose regimens, we also sought to investigate the effect of cumulative aspirin dose on AD risk. The inconsistent recording of medication end dates, dosing frequency (e.g., daily versus as needed), and therapy duration within the EHR, along with multiple sources of medication documentation in patient charts (including medication lists in clinical notes as well as prescriptions), hindered precise quantification of total aspirin exposure using EHR data. To address this limitation, we developed a proxy measure: the documented aspirin exposure rate, defined as the total number of unique aspirin records divided by the time (in years) between the first and last recorded aspirin exposures. This measure was intended to capture the frequency and duration of aspirin use documented in the EHR, with a higher rate reflecting more consistent and sustained aspirin exposure. In a matched analysis accounting for sex, race, baseline comorbidities (cardiovascular disease, cerebrovascular disease, and rheumatoid arthritis), EHR time after age 65, number of EHR visits, and aspirin exposure duration, we observed that patients with an aspirin exposure rate above the median (>5 exposures per year) had a 42% lower risk of AD compared to those below the median (HR=0.58, 95% CI: 0.39-0.88, *P*=0.009).

To assess the generalizability of these findings, we performed two sets of external replication studies in which we independently evaluated the relationship between aspirin and AD, using (1) the NIH *All of Us* Research Program database and (2) the combined MarketScan Commercial Claims and Encounters and Medicare Supplemental databases. We were able to use the same retrospective cohort study design with survival analysis to investigate the association between aspirin exposure and AD using EHR data from *All of Us*; however, we did not have adequate follow-up time to capture AD-relevant endpoints in survival analysis using the MarketScan claims data. In MarketScan, the mean age at maximum follow-up among individuals exposed to aspirin before age 65 was 75.8 ± 8.2, whereas the mean age of first AD diagnosis in the overall patient population was 84.2 ± 7.2. Thus, to investigate the association between aspirin use and AD in MarketScan, we performed a case-control study in which we captured AD outcomes first and then ascertained previous aspirin exposure.

Our analysis using *All of Us* EHR data confirmed a protective effect for aspirin against AD (HR=0.40, 95% CI: 0.15-1.08, *P*=0.07), although these results were limited by low statistical power, with only 24 total AD events observed among the entire cohort of 1,995 patients. We performed a meta-analysis combining the hazard ratios obtained from VUMC and *All of Us*. In the meta-analysis, we observed that aspirin use before age 65 was associated with 24% decreased incidence of AD (HR=0.76, 95% CI: 0.64-0.89, *P*=0.001). In the case-control study using claims data from MarketScan, we found that AD was associated with significantly lower odds of prior aspirin exposure (OR=0.32, 95% CI: 0.28-0.38, *P*<0.001), indicating that patients diagnosed with AD were less likely to have been previously exposed to aspirin. Due to the limited number of AD events in the *All of Us* cohort and the small number of aspirin prescriptions in the MarketScan cohort, we were unable to conduct meaningful analyses investigating the effects of aspirin dosage or exposure rate in these datasets.

**Fig. 3:**
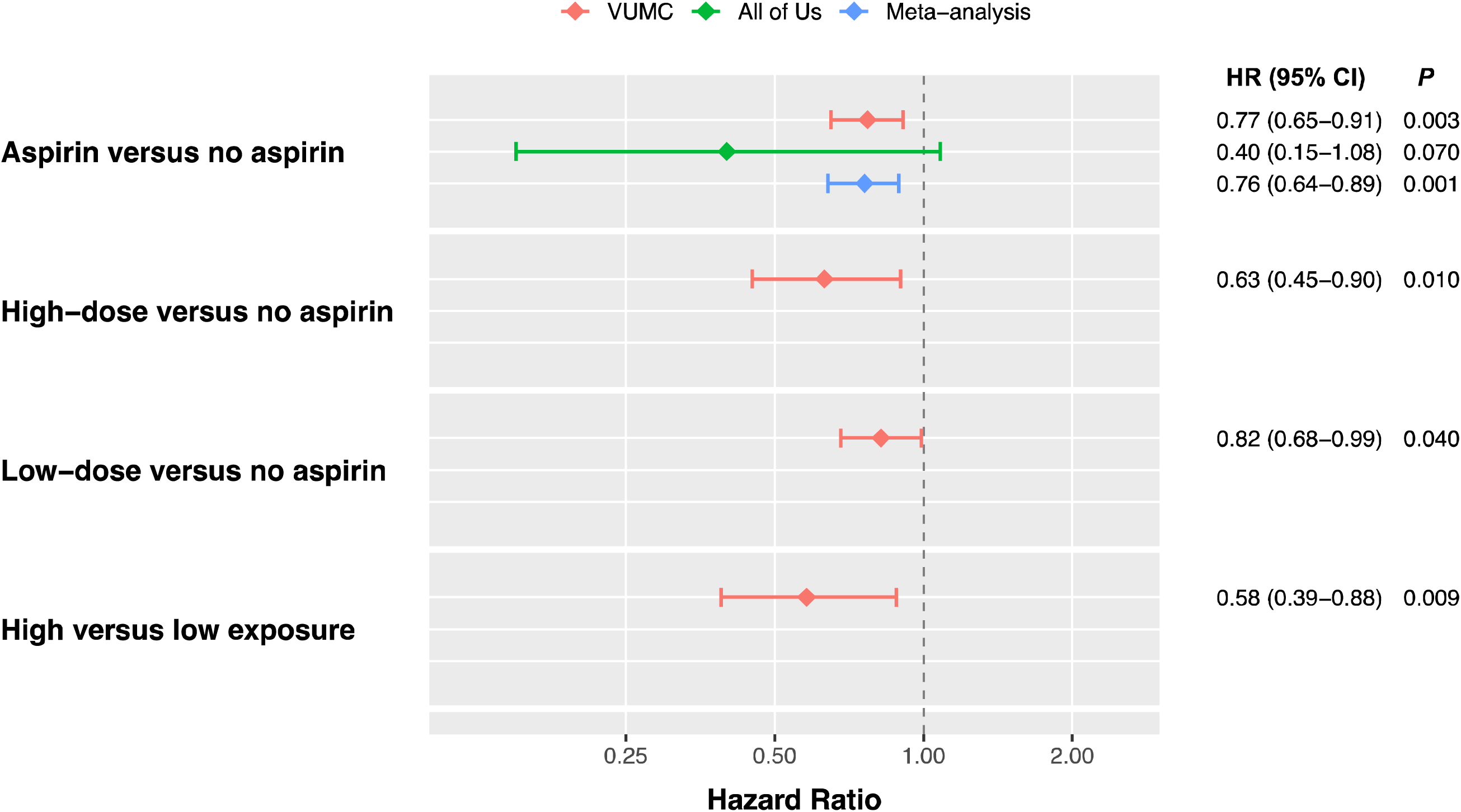
Associations between aspirin use before age 65 and risk of incident AD. From top to bottom, HRs and 95% CIs are shown for: (1) any aspirin use compared to no aspirin use, (2) high-dose (≥325mg/day) compared to no aspirin, (3) low-dose (≤81mg/day) compared to no aspirin, and (4) high aspirin exposure rate (greater than the median rate of 5 documented exposures per year) compared to low exposure rate. Results are displayed separately for VUMC, *All of Us*, and the combined meta-analysis. AD Alzheimer’s disease, HR hazard ratio, CI confidence interval.

In addition, we conducted stratified analyses based on *APOE* genotype using data from VUMC and *All of Us* (the MarketScan Research Databases do not contain genetic data). Of the 19,413 patients in the VUMC study cohort, 1,856 patients had genetic data available for *APOE* in VUMC’s EHR-linked DNA biobank, with a total of 471 *APOE* ε4 carriers (*N*=41 homozygous carriers, *N*=430 heterozygous carriers). The *All of Us* cohort contained 1,450 patients genotyped for *APOE*, with 294 *APOE* ε4 carriers (*N*=23 homozygous, *N*=271 heterozygous). Among patients with at least one copy of the *APOE* ε4 allele at VUMC, aspirin use before age 65 was associated with a 40% decreased risk of incident AD after age 65, although this effect was not statistically significant (HR=0.60, 95% CI: 0.33-1.10, *P*=0.0986). In *All of Us*, aspirin use was associated with a 52% reduced risk of AD (HR=0.48, 95% CI: 0.10-2.25, *P*=0.35) among patients with at least one copy of the *APOE* ε4 allele, although this effect was not statistically significant. In meta-analysis combining these findings from VUMC and *All of U*s, aspirin use before age 65 was associated with a 41% decreased risk of incident AD after age 65 (HR=0.59, 95% CI: 0.33-1.02, *P*=0.06) in patients with at least one copy of the *APOE* ε4 allele, suggesting a possible protective effect in this patient population in the context of limited statistical power. In contrast, aspirin use was not significantly associated with AD among patients without any copies of the *APOE* ε4 allele (VUMC HR=0.63, 95% CI: 0.26-1.55, *P*=0.317; *All of Us* HR=0.52, 95% CI: 0.15-1.79, *P*=0.3; meta-analysis HR=0.59, 95% CI: 0.29-1.22, *P*=0.16), although the power of these analyses was limited by the small number of AD cases.

## Discussion

In this study, we identified aspirin, a widely used non-steroidal anti-inflammatory drug (NSAID), as a promising repurposing candidate for the prevention of AD. The study utilized publicly available multi-omics data to generate drug repurposing hypotheses and real-world data in the form of electronic health records and health insurance claims to clinically validate the effects of aspirin on AD risk. Aspirin emerged as a candidate based on a transcriptomic analysis that suggested it could reverse AD-associated changes in gene expression in bulk brain and peripheral tissues as well as in microglia. Using two independent EHR datasets, we then demonstrated an association between aspirin use before age 65 and reduced risk of incident AD, strengthening our initial drug repurposing hypothesis. Analysis of national healthcare claims data further corroborated these findings, showing that individuals with AD were less likely to have previous aspirin prescriptions.

We generated transcriptomic signatures of AD using eQTL data from both the GTEx project and Microglia Genomic Atlas (MiGA). Our disease signatures recapitulated several findings from other studies. In all three of the GTEx-derived AD signatures, we observed inverse associations of *TOMM40* (*Z*_brain_ = -8.05, *Z*_AD-relevant_ = -7.90, and *Z*_all_ = -6.75) and *PVR* (*Z*_brain_ = -3.15, *Z*_AD-relevant_ = -3.15, and *Z*_all_ = -3.77) with AD risk, consistent with previous gene-based associations identified in the dorsolateral prefrontal cortex using data from the Common Mind Consortium (*TOMM40 Z* = -21.40, *PVR Z* = -4.82)^27^. Additionally, we observed an inverse association of *CR1* with AD risk in the brain-tissue GTEx signature, a positive association of *DMWD* with AD risk in the AD-relevant GTEx signature, as well as positive associations of *IRF2BP1* and *DMWD* and an inverse association of *CLPTM1* with AD risk in the all-tissue GTEx signature, which have been reported previously^27^. We also identified several known AD-related changes in gene expression within our microglial signature. Notably, we observed a positive association of *CD33* expression with AD risk in our microglial AD signature, which is consistent with the findings of previous studies of microglia in AD^37,62^. We also observed positive disease associations for several major histocompatibility complex (MHC) class II genes expressed by activated microglia, including *HLA-DRB1*, *HLA-DRB5*, and *HLA-DQA1*, which have been associated with AD in other studies^38,63^.

While the GTEx signatures captured bulk tissue gene expression encompassing multiple cell types, the microglial signature enabled us to focus selectively on AD-associated gene expression in one of the leading cell types implicated in the disease. In recent years, microglia have been found to play a key role in the pathogenesis of AD through multiple mechanisms, including mediation of neuroinflammation, regulation of amyloid-β clearance, synaptic pruning, and maintenance of neuronal homeostasis^64^. Microglia have also been observed to undergo unique transcriptional changes in AD^65^, suggesting the utility of generating microglia-specific gene expression signatures of AD to identify possible AD drug repurposing candidates. Our results revealed significant variation in genetically regulated gene expression in microglial cells compared to bulk tissues, even bulk brain tissues. Notably, we observed minimal overlap in the genes encompassed by the GTEx AD signatures and the microglia AD signature—only nine AD risk genes were shared by all four signatures, and out of these nine genes, only seven showed concordant effect directionalities. These findings reflect the importance of leveraging both bulk and cell type-specific transcriptomic data to identify gene-disease associations, particularly for complex diseases such as AD. Certain AD risk genes, and thus possible therapeutic targets, may be overlooked when examining one tissue context or averaging signals across heterogeneous cell populations. Furthermore, the contrasting gene expression associations with AD risk observed in the GTEx and microglial signatures may reflect distinct regulatory mechanisms and functional roles in different cellular environments. For example, while *BIN1* was positively associated with AD risk in the microglial AD gene expression signature, it was inversely associated with AD risk in all three GTEx-derived disease signatures. Previous studies have suggested that increased *BIN1* expression within microglia may contribute to AD pathogenesis by modulating tau protein aggregation and clearance^66,67^; however, studies involving other cell types have observed decreased expression of *BIN1* in AD^68,69^.

Among the compounds exhibiting negative connectivity to AD gene expression changes in our CMap queries, we focused our real-world validation studies on aspirin, given its widespread use in clinical care for primary and secondary prevention of cardiovascular disease and its well-established safety profile, making it an accessible potential treatment option for AD as well as a favorable candidate for investigation in EHR and claims data. From a mechanistic standpoint, aspirin presents an intriguing drug repurposing candidate for AD. Studies have suggested that chronic activation of inflammatory processes may play a key role in AD pathogenesis and neurodegeneration^70^. Activated microglia have been found to surround amyloid plaques in brains affected by AD^71^, and elevated levels of inflammatory markers have been found in the brains and cerebrospinal fluid of patients with AD^72,73^. By inhibiting cyclooxygenase (COX) enzymes, aspirin inhibits platelet aggregation and reduces the synthesis of pro-inflammatory prostaglandins, which are thought to contribute to the chronic neuroinflammation observed in AD^74^.

In our clinical validation studies, we observed that aspirin use before age 65 was associated with a 24% reduced risk of incident AD. More detailed analysis suggested that the protective effect of aspirin on AD risk may depend on cumulative exposure rather than dosing strength alone. While we were not able to detect a clear difference between high-dose and low-dose aspirin regimens using EHR data, we found that individuals with a higher documented aspirin exposure rate had a significantly lower risk of incident AD compared to matched individuals with lower exposure rates. These findings suggest that greater cumulative aspirin exposure, reflecting either a higher total aspirin amount or more sustained aspirin use over time, may be necessary to confer a protective benefit against AD.

Although aspirin has been previously investigated in AD, studies have yielded mixed results. While early observational studies suggested that aspirin might reduce the risk of developing AD^75^, randomized clinical trials such as ASPREE^76^ and AD2000^77^ did not show a clear protective benefit for aspirin in reducing cognitive decline. Several important differences may explain these inconsistent findings. First, the timing of aspirin exposure varied significantly across studies, as well as the duration of follow-up. The ASPREE trial investigated 100 mg daily aspirin initiated in healthy participants aged ≥70 years (≥65 for Hispanics and African Americans in the United States), with a median follow-up of 4.7 years^76^. However, our study focused on early aspirin exposure, with first aspirin use occurring before age 65. Importantly, in our study, we observed that the mean age of first AD diagnosis was ∼76 years in both VUMC and *All of Us*, suggesting that less than 5 years of follow-up after ages 65 to 70 may be insufficient to assess the effects of aspirin on a slowly progressive disease like AD. In contrast, the AD2000 trial evaluated the effects of 75 mg daily aspirin in participants with pre-existing cognitive decline, with a median age of 75 years^77^, assessing aspirin primarily as a treatment for clinical, symptomatic AD rather than as a preventive measure or treatment for preclinical AD.

The strengths of this study lie in the use of a multi-omics approach to suggest drug repurposing hypotheses for AD combined with real-world validation leveraging clinical data from multiple independent databases. While previous studies aimed at identifying drug repurposing candidates for AD have suggested a variety of compounds, including antidiabetic medications (i.e., metformin^23^), antihypertensives (e.g., angiotensin receptor blockers^23,78^, bumetanide^17^), and sildenafil^18^, a major challenge in drug repurposing research has been a lack of validation across diverse clinical datasets. Real-world data has been utilized infrequently to confirm preliminary drug repurposing signals^79^. One of the main strengths of our study is the ability to follow patients longitudinally for extended periods of time, capturing timepoints and outcomes relevant to AD that may be missed in short-term clinical trials. Furthermore, by using EHR data, we were able to investigate the effects of early aspirin initiation (i.e., first aspirin use before the age of 65) on AD risk decades later, which most randomized trials are not able to assess. We were also able to perform deeper phenotyping to investigate differences in AD incidence based on high-dose and low-dose aspirin use, as well as high versus low documented aspirin exposure rates (intended to serve as a proxy measure of cumulative dose). We identified a protective effect for aspirin against AD in two large EHR databases, VUMC and *All of Us*. While we were not able to replicate the survival analysis using health insurance claims data due to the shorter observation periods available in the MarketScan dataset, we were able to confirm a protective effect for aspirin using a case-control study design. Additionally, we leveraged genetic data from VUMC and *All of Us* to explore potential differences in aspirin’s effects on AD risk between individuals with and without at least one copy of the *APOE* ε4 allele, a genetic variant known to markedly increase AD risk^55^. Although the relatively small number of *APOE* ε4 carriers as well as the low prevalence of AD diagnoses among *APOE* ε4 non-carriers restricted the statistical power of our analyses, our findings nonetheless suggest a possible protective effect for aspirin specifically for *APOE* ε4 carriers. These results demonstrate the potential for EHRs and linked biobanks to support precision drug repurposing initiatives aimed at identifying patient subgroups most likely to benefit from a particular therapeutic intervention, particularly as the amount of clinical and genetic data contained in these resources continues to grow.

This study has several limitations. First, the AD drug repurposing hypotheses we generated were limited to the small-molecule compounds tested in the CMap database, which are not exhaustive and do not represent all possible repurposing opportunities for AD. Furthermore, the drugs in CMap have been primarily tested in non-brain cancer cell lines, which may not precisely reflect gene expression patterns in AD-relevant cell types such as microglia and neuronal cells. Second, in constructing the AD gene expression signatures used to query CMap, we were limited in population diversity. Most existing GWAS have been conducted among individuals of European ancestry, which may limit the generalizability of the gene expression signatures and drug repurposing candidates identified using such GWAS. In addition, although AD incidence differs among populations^80^, we did not have sufficient power in these cohorts to perform meaningful race-stratified analyses. Notably, while African American and Hispanic patients comprise ∼14% of the EHR in BioVU, they represented <8% of the qualifying cohort. Similarly, although *All of Us* includes ∼45% racial minorities, only 7% of the qualifying cohort was African American; further, race data is not reported in the MarketScan databases. Third, we relied on structured diagnosis codes to identify AD patients. While useful for large-scale studies, this approach does not encompass the full clinical heterogeneity of AD, including disease severity and progression. The accuracy of coding is also a concern, as AD may be inappropriately attributed to other causes of dementia. However, to mitigate the risk of outcome misclassification, we explicitly excluded individuals with any diagnosis of non-AD dementia occurring at any time. Importantly, any residual misclassification was unlikely to be dependent on exposure and would affect both the aspirin-exposed and unexposed cohorts similarly. Fourth, we acknowledge that aspirin use may not be accurately documented for all patients in the EHR given its over the counter availability. Although documentation is likely more complete for patients with a history of cardiovascular disease or stroke, aspirin taken daily for general health purposes may be underreported in the EHR. Nevertheless, conducting analysis limited to these documented records may better identify systemic, rather than incidental, aspirin use (as would be the case if all over the counter usage was included). Similarly, capturing aspirin use in claims data presents challenges, as the MarketScan dataset includes only filled prescription claims. Consequently, if a physician advised over-the-counter aspirin, such usage would not appear in the outpatient drug files—a limitation that may explain the low number of aspirin prescriptions observed in the MarketScan data.

In summary, in this study, we identified aspirin as a drug repurposing candidate for AD using a virtual transcriptome approach to systematically screen for compounds capable of reversing AD-associated changes in gene expression. To increase confidence in our drug repurposing hypothesis, we then analyzed the relationship between aspirin use and AD in multiple clinical datasets. Our results using EHR data from VUMC and *All of Us* suggest the benefits of early initiation of aspirin (before the age of 65) to prevent AD later in life. These findings provide a rationale for designing clinical trials to further evaluate aspirin as a preventive agent. Future studies should investigate whether early aspirin initiation can delay or reduce the onset of AD, while also determining the optimal dosing regimens required to maximize neuroprotective benefits and minimize potential adverse effects. As data continues to accumulate, EHRs can be expected to play an increasingly important role in informing the development of evidence-based strategies and precision medicine approaches for both preventing and treating AD.

## Methods

This study was conducted with approval from the VUMC Institutional Review Board and the NIH *All of Us* Research Program. All EHR data from VUMC and *All of Us* are de-identified; use of these data is considered non-human subjects research.

### Construction of microglia transcriptome prediction models

We downloaded full nominal eQTL summary statistics for 255 primary human microglia samples collected from four different regions of the brain from 100 human subjects from the Microglia Genomic Atlas (MiGA)^29^. These four brain regions encompassed two cortical areas, the medial frontal gyrus and superior temporal gyrus, and two subcortical areas, the subventricular zone and thalamus. Details on genotyping, RNA-seq generation and processing, and eQTL mapping can be found in the MiGA flagship paper^81^. We followed the multivariate adaptive shrinkage approach to eQTL analysis introduced by Urbut et al.^82^ to build microglia-specific Multivariate Adaptive Shrinkage in R (MASHR) transcriptome prediction models for each brain region. We applied two different approaches to determine which SNPs would be included in the final MASHR models. First, we focused only on the strongest cis-eQTL signals, selecting only the top eQTL (with the largest univariate |Z|-statistic across the four brain regions) per gene for the final models (*N* = 18,799 SNPs in total). However, we found that there was very limited overlap between the SNPs in these MASHR models and the SNPs in the AD GWAS summary statistics used to run S-PrediXcan and S-MultiXcan; therefore, we also built a separate set of microglia MASHR models including more eQTLs. In this second approach, for each brain region, we selected the top five eQTLs with the lowest local false sign rate (a measure analogous to false discovery rate)^82^ per gene and included these eQTLs in the final models. In cases where multiple eQTLs were tied for lowest local false sign rate, we retained all eQTLs.

### Imputation of transcriptomic signatures for Alzheimer’s disease

We applied two statistical methods, S-PrediXcan^24^ and S-MultiXcan^25^, to publicly available Alzheimer’s GWAS summary statistics to compute transcriptomic signatures for AD. S-PrediXcan and S-MultiXcan predict gene expression for a trait or disease of interest using prediction models trained on reference transcriptome datasets. S-PrediXcan computes single-tissue gene-level association results from GWAS summary statistics. S-MultiXcan then aggregates the single-tissue S-PrediXcan results across multiple tissues, thereby increasing the statistical power to detect associations. We used GWAS summary statistics for a total of 762,917 individuals (86,531 AD cases and 676,386 controls, excluding participants from 23andMe)^26^ to generate all AD transcriptomic signatures used in this study. To improve GWAS-QTL integration, we harmonized the AD GWAS summary statistics to GTEx v8 variants and imputed summary statistics for missing variants, as described in the MetaXcan best practices guide^83^.

We first used S-PrediXcan to impute single-tissue gene expression levels in 49 available GTEx tissues, using pre-developed MASHR expression prediction models^24,84,85^. We then ran S-MultiXcan on the S-PrediXcan results to predict gene expression across multiple tissues. Although AD predominantly affects the brain, studies have suggested that peripheral tissues such as the skin and vascular tissues can also capture genetic effects on gene expression in AD and may even represent potential pathogenic tissues in AD^27^. Thus, we conducted three separate S-MultiXcan analyses to predict AD-associated changes in gene expression combining brain and non-brain tissues, including: (1) a brain-specific analysis combining predictions from the 13 GTEx brain tissues, (2) an AD-relevant tissue analysis combining the 13 brain tissues with four other tissues previously related to AD (whole blood, spleen, and two skin tissues)^27,28^, and (3) a non-specific analysis combining predictions from all 49 GTEx tissues. We then constructed a separate AD virtual transcriptomic signature for each of the brain-restricted, AD-relevant, and all-tissue S-MultiXcan analyses using the AD risk genes with ≥2/3 tissue concordance (i.e., showing the same directionality of gene expression changes in at least two-thirds of the tissues with non-NA results). We defined AD risk genes using Bonferroni correction (*P*<0.05/number of S-MultiXcan gene associations). We used the mean *Z* score among the concordant tissues to classify the risk genes as positively associated with AD risk (mean *Z* score>0) or inversely associated with AD risk (mean *Z* score<0)

We also used S-PrediXcan and S-MultiXcan to calculate a microglia-specific AD gene expression signature, using the larger microglia MASHR transcriptome prediction models we trained with MiGA data in our second approach. To run S-PrediXcan with our custom models, we needed to calculate covariance matrices to account for linkage disequilibrium (LD) between SNPs. We used reference data from the 1000 Genomes Project^86^ to generate these covariance matrices. We then applied S-PrediXcan to predict microglial gene expression in the medial frontal gyrus, superior temporal gyrus, subventricular zone, and thalamus. We initially used S-MultiXcan to construct a single AD gene expression signature integrating the S-PrediXcan results for microglia in the four different brain regions; however, the high correlations between the S-PrediXcan associations across the brain regions prevented computation of S-MultiXcan association statistics for the majority (>80%) of the genes tested. We thus used a generalized Berk-Jones (GBJ) test to combine the S-PrediXcan *Z* scores from the four brain regions^30^. Again, AD risk genes were defined using a Bonferroni-corrected significance threshold (GBJ *P*<0.05/18,222 genes tested). The GBJ test does not provide directional effects (i.e., the calculated GBJ statistic is always positive). To account for this, we included only the AD risk genes with concordant effect directions across all brain regions with available results in the microglial AD virtual transcriptomic signature. NA values did not influence the concordance assessment. To capture inverse associations with AD risk, we negated the GBJ statistic for genes with consistently negative S-PrediXcan *Z* scores across the brain regions.

### Identification of Alzheimer’s disease drug repurposing candidates

We queried the Connectivity Map (CMap) drug perturbation database to identify drugs that could reverse the AD gene expression signatures imputed by S-MultiXcan. CMap has created a library with over 1.5 million gene expression profiles (including ∼1,000 directly measured landmark genes) capturing the effects of over 8,000 small-molecule compounds and genetic reagents in a variety of human cell lines^21^. A key feature of CMap is its ability to compare disease gene expression signatures against the perturbagen-induced signatures contained in its library, facilitating the discovery of drugs that could reverse or mimic a particular disease state. To identify AD drug repurposing candidates, we uploaded a list of the identified risk genes in each AD transcriptomic signature to the CMap CLUE platform, which we used to query the CMap database. CMap computes a connectivity score that quantifies the similarity between the queried signature and each perturbagen signature. We defined potential drug repurposing candidates as those small-molecule compounds with negative connectivity to the input AD gene expression signature, as has been done in previous drug repurposing studies^14,17^.

### Clinical validation using real-world data

To validate our genetic drug repurposing hypothesis suggesting aspirin as a potential AD therapeutic, we investigated the effects of aspirin exposure in humans using clinical data from electronic health records (EHRs) and health insurance claims. In the two EHR databases, we performed a retrospective cohort study evaluating the risk of new AD diagnosis after age 65 in individuals previously exposed to aspirin compared to unexposed individuals to determine whether aspirin was associated with lower incidence of AD. In the claims database, we performed a case-control study comparing the odds of aspirin use in AD cases and controls.

#### Data sources

We performed clinical validation studies using diagnosis and medication data from three independent sources: (1) VUMC’s Synthetic Derivative (SD) database, (2) the NIH *All of Us* Research Program database, and (3) the MarketScan Commercial Claims and Encounters (CCAE) and Medicare Supplemental (MDCR) databases. The VUMC SD contains decades of longitudinal clinical data, including diagnosis and procedure codes, laboratory test results, and medications, extracted from de-identified EHRs for over 3.4 million unique patients^87^. The SD is linked to VUMC’s biobank, BioVU, which contains over 300,000 unique DNA samples as of January 2023, allowing for the integration of EHR and genetic data. At the time of the study, the *All of Us* Research Program database contained data for over 633,540 participants, with genomic sequencing data for over 414,840 participants^88^. The MarketScan CCAE and MDCR databases collectively contain detailed insurance claims data, including diagnosis codes and pharmacy claims, for over 200 million unique patients.

#### Cohort formation and outcome assessment: VUMC and All of Us

To capture endpoints relevant to AD and ensure adequate EHR follow-up time was available for all patients in the study population, we restricted our analysis to patients over 65 years of age with at least one visit at ≥75 years with no prior diagnosis of AD or dementia. We excluded individuals with AD diagnosed at ≤65 years, as well as individuals with diagnoses of non-AD dementia (occurring at any time).

Information on patient medications is available as structured data in the VUMC and *All of Us* EHR databases (within a DRUG_EXPOSURE table, in accordance with the Observational Medical Outcomes Partnership Common Data Model^89^). Drug exposures documented in this table may originate from various sources in the EHR, including clinical notes (e.g., medication lists appearing in history and physical (H&P) reports, progress notes, and discharge summaries), inpatient and outpatient medication orders (e.g., prescriptions), and historical medication records.

We identified aspirin-exposed patients by mapping patient medications to their ingredients using the RxNorm standardized terminology for clinical drugs and filtering for all medications containing aspirin (RxCUI = 1191). We used aspirin-exposed patients with at least one year of documented aspirin use prior to age 65 to form the aspirin-exposed group; patients without any documented aspirin use were considered unexposed controls. Patients with first aspirin use occurring after age 65 were excluded. We then identified two groups within the aspirin-exposed cohort for additional comparisons: high-dose (≥325mg/day) and low-dose (≤81mg/day) aspirin users, excluding individuals exposed to intermediate aspirin doses and individuals switching between low-dose and high-dose aspirin use.

Aspirin is used in the treatment of multiple medical conditions, including atherosclerotic cardiovascular disease (in low doses for primary prevention) and inflammatory diseases such as rheumatoid arthritis (in high doses for pain relief), which may also impact AD risk. To minimize the effects of confounding by indication and other sources of confounding, we matched patients in the aspirin-exposed cohort to control patients with no recorded aspirin use in a 1:2 ratio, using a propensity score based on sex, race, EHR time after age 65, and presence of clinical indications for aspirin use (cardiovascular disease, cerebrovascular disease, and rheumatoid arthritis) at baseline (defined as the time 0 of age 65 or, if no data available at age 65, the age of the first EHR visit). We used the MatchIt R^90^ package to perform nearest-neighbor propensity score matching. The diagnoses used to define the comorbidities for matching are provided in **Supplementary Tables 11**-**13**.

We defined AD cases using a requirement of at least one of the following AD diagnosis codes: ICD-9-CM code 331.0 or ICD-10-CM codes G30.1, G30.8, and G30.9. We excluded patients diagnosed with AD before age 65, as well as patients diagnosed with other forms of dementia (vascular dementia, diffuse Lewy body disease, frontotemporal dementia, mixed dementia, and dementia associated with Parkinson’s disease) (**Supplementary Table 14**).

#### Statistical analysis: VUMC and All of Us

We used Cox proportional hazards regression models to investigate the risk of AD after age 65 in aspirin-exposed and unexposed individuals. First, we compared AD risk between the aspirin-exposed cohort and the propensity score-matched unexposed cohort. Next, we performed subgroup analyses based on aspirin dose (high-dose versus low-dose versus no aspirin), documented aspirin exposure rate, and *APOE* ε4 genotype.

We used the metafor R package for meta-analysis of hazard ratios^91^. We used Cochran’s Q test and the I^2^ index to assess heterogeneity. Based on these measures of heterogeneity, all meta-analyses were conducted under a fixed-effects model.

#### Documented aspirin exposure rate

We also evaluated the relationship between the documented aspirin exposure rate and AD risk among aspirin users. We calculated the documented aspirin exposure rate for each individual in the aspirin-exposed cohort by dividing the number of unique aspirin exposures recorded in their EHR by the duration (in years) between their first and last documented aspirin exposures. We then classified aspirin users into high- and low-exposure groups based on the median documented exposure rate of 5 aspirin exposures per year. To ensure balanced comparisons, we matched individuals with high documented aspirin exposure rates to those with low rates in a 1:1 ratio using propensity score matching. The variables used in matching were sex, race, baseline comorbidities (cardiovascular disease, cerebrovascular disease, rheumatoid arthritis), EHR time after age 65, total number of EHR visits, and aspirin duration. The final matched cohort comprised 3,690 individuals (*N* = 1,845 per group). A Cox proportional hazards regression model was used to investigate the risk of AD after age 65 in the high-exposure group relative to the low-exposure group.

#### APOE genotyping

*APOE* genotype was determined using the combination of alleles at SNPs rs429358 and rs7412. The *APOE* ε4 variant was defined as the presence of a C allele at both SNPs. Genetic data was available for 1,856 patients in the VUMC cohort (including 41 APOE ε4 homozygotes and 430 heterozygotes) and 1,450 patients in the *All of Us* cohort (with 23 APOE ε4 homozygotes and 271 heterozygotes). Information on *APOE* genotype was not available in the MarketScan database.

#### MarketScan validation

Given the shorter observation time in the MarketScan Research Databases, which prevented us from capturing AD-relevant timepoints in patients exposed to aspirin before age 65, we performed a case-control study to investigate the association between aspirin use and AD. We first identified AD cases using ICD-9-CM code 331.0 and ICD-10-CM codes G30.1, G30.8, and G30.9. We matched AD cases to comparable controls in a 1:2 ratio based on propensity score, using sex, comorbidities (cardiovascular disease, cerebrovascular disease, and rheumatoid arthritis, diagnosed at any age), and claims follow-up time (difference in years between first and last claims records) as covariates. We did not match on race as this is not reported in MarketScan. Aspirin prescriptions were identified using National Drug Codes. We then calculated the odds ratio for aspirin exposure among the AD cases compared to their matched controls.

## Data Availability

All data generated or analyzed during this study are included in this published article and its Supplementary Information. The microglia eQTL summary statistics from the Microglia Genomic Atlas used in this study can be downloaded from the NIAGADS Data Sharing Service using accession number NG00105.v3. The AD GWAS summary statistics used in this study are available at https://cncr.nl/research/summary_statistics/. The MASHR GTEx v8 transcriptome prediction models can be downloaded from PredictDB (https://predictdb.org/categories/downloads/). Access to VUMC EHR’s database requires institutional approval and compliance with a data use agreement. Data from the *All of Us* Research Program can be accessed through the Researcher Workbench (https://workbench.researchallofus.org). The MarketScan claims data used in this study can be requested from Merative®.

## Acknowledgements

This study was supported by the National Institute of Aging of the National Institutes of Health under award numbers R01AG069900, F30AG080885.

---

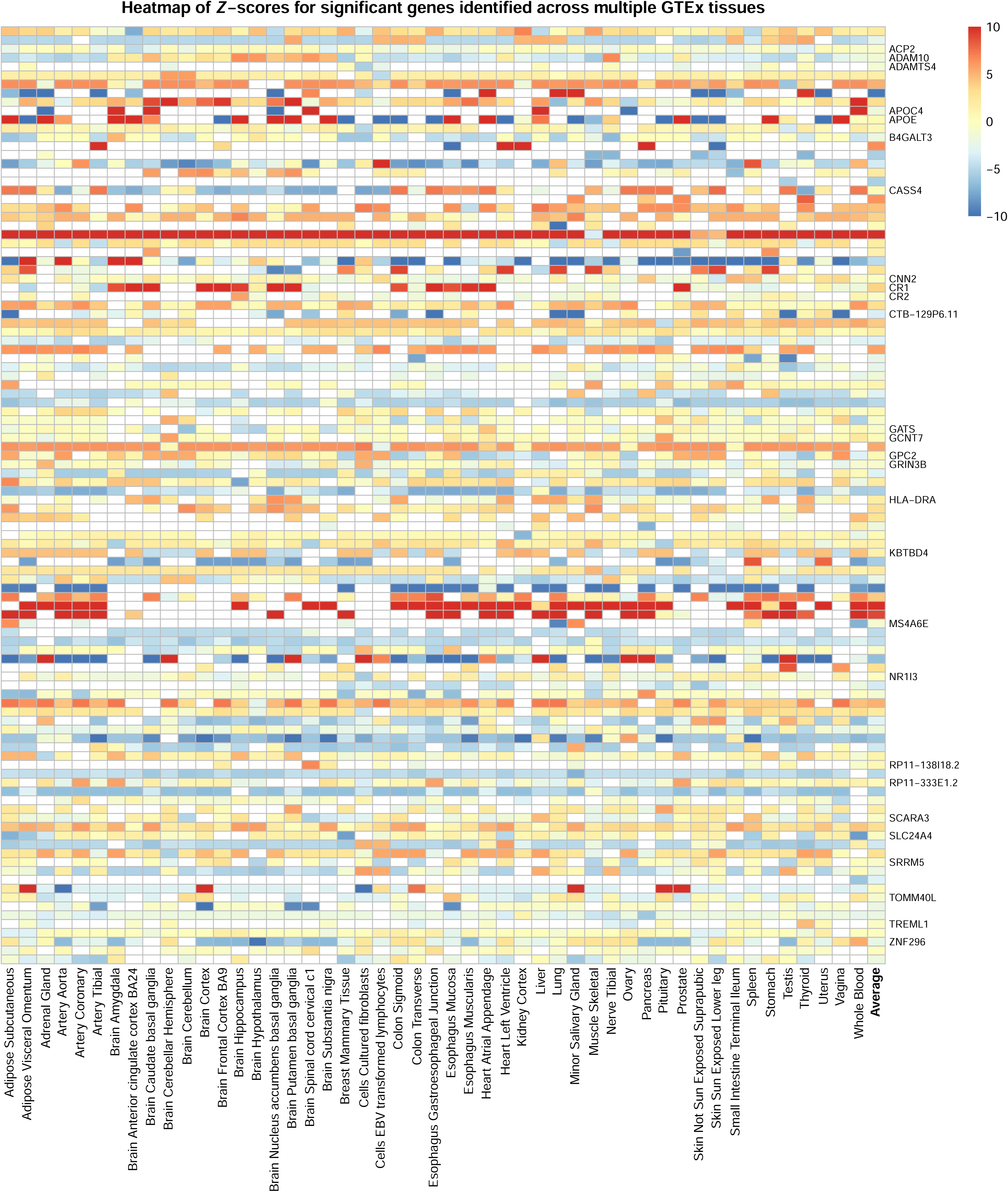

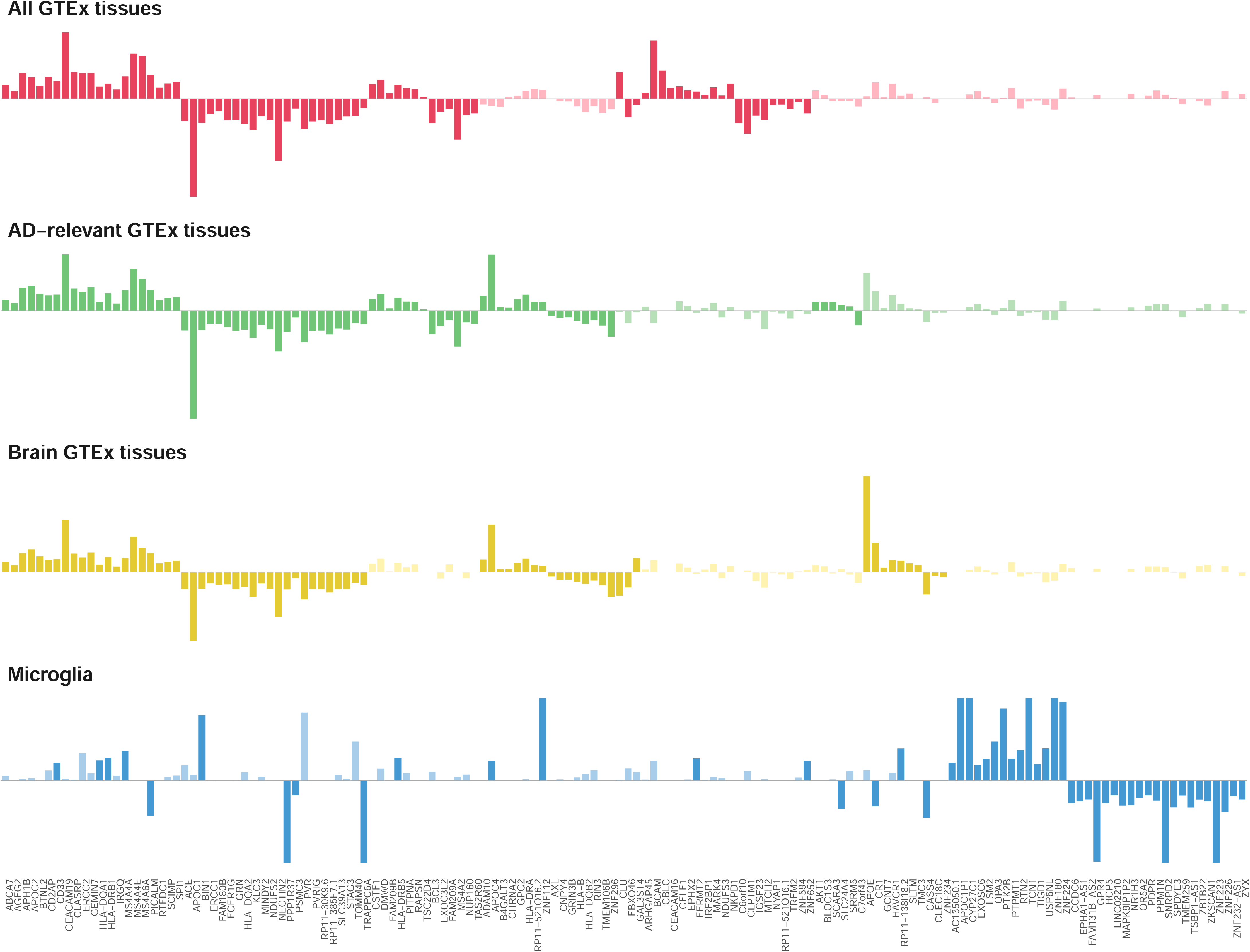

